# Clinical Assessment of a Plasma AT(N) Panel for Alzheimer’s Disease

**DOI:** 10.1101/2024.08.06.24310938

**Authors:** Bradley B. Collier, Matthew R. Chappell, Whitney C. Brandon, Tien Le, Ayla B. Harris, Joseph M. Volpe, Robert Martone, John W. Winslow, Deborah Boles, Andre Valcour, Russell P. Grant

## Abstract

1.

**Importance:** With the emergence of new therapeutics for treatment of Alzheimer’s disease, there is currently a critical need for sensitive and accurate blood-based tests to assist with the diagnosis and treatment of Alzheimer’s disease.

**Objective:** To determine the clinical validity of an analytically validated plasma panel for the assessment of Alzheimer’s disease.

**Design, Setting, and Participants:** This cross-sectional study measured biomarkers representative of the Alzheimer’s disease AT(N) framework in 200 plasma specimens acquired from the Australian Imaging, Biomarker & Lifestyle (AIBL) Study of Ageing. Specimens were obtained from amyloid PET negative subjects classified as cognitively unimpaired (n = 75) and amyloid PET positive subjects classified as having no cognitive impairment (n = 49), mild cognitive impairment (n = 26), or Alzheimer’s disease dementia (n = 50).

**Exposures:** Amyloid PET and plasma Aβ42/40, pTau181, and NfL.

**Main Outcomes and Measures:** To assess the utility of the plasma panel to assess onset and progression of Alzheimer’s disease with respect to amyloid PET results and cognitive impairment.

**Results:** A difference was observed for each assay with respect to amyloid status (p<0.0001). Receiver operating characteristic (ROC) analysis of clinical specimen results from validated assays produced an area-under-the-curve (AUC) of 0.941 for Aβ42/40, 0.847 for pTau181, and 0.666 for NfL (p < 0.0001 for all biomarkers). The sensitivity (96.0%) and specificity (86.7%) observed for Aβ42/40 measurements meets current recommendations for triage testing. In addition, plasma levels of pTau181 and NfL were also found to increase with worsening cognitive impairment.

**Conclusions and Relevance:** The clinical concordance with amyloid PET for each biomarker is consistent with the biological progression of the AD continuum. As such, the availability of this AT(N) panel will provide clinicians with a simple blood-based means to provide evidence of AD pathological changes and could help identify AD patients much faster, shorten the overall AD patient diagnostic journey, and enable earlier treatment interventions.

## 2. Introduction

Alzheimer’s disease (AD) is understood to be a progressive neurodegenerative disease that presents as a continuum from a presymptomatic/prodromal stage to severe dementia.^1–6^ In conjunction with this understanding, a biomarker framework was proposed for assessing the underlying brain pathology of AD irrespective of patient symptoms.^1–6^ The AT(N) framework consists of various biomarkers traditionally grouped into three categories based on the type of Alzheimer’s related pathology investigated:

- **A** – biomarkers associated with amyloid pathology (*e.g.* plaques)
- **T** – biomarkers associated with tau pathology (*e.g.* neurofibrillary tangles)
- **(N)** – biomarkers associated with neurodegeneration

where the order listed is representative of the typical progression of AD and *N* is placed in parentheses to indicate that neurodegeneration is not specific to AD.^2,6–8^

Positron emission tomography (PET) and other imaging techniques as well as measurements of cerebrospinal fluid (CSF) biomarkers have demonstrated utility for identifying Alzheimer’s disease (AD) pathology in each of the three AT(N) categories; however, these tools remain costly, invasive, and/or not widely available.^9–12^ Due to the growing prevalence of this progressive disease and emerging therapeutics, robust blood plasma-based measurements are a desirable alternative.^3,9,11–13^

Advancements in plasma biomarker platforms, which enable more accurate identification of disease pathology at lower levels of detection, led to the extension of the AT(N) framework to support the clinical diagnosis of AD.^5,7,14^ The AT(N) framework provides an objective approach to determine the status of biological changes that are indicative of Alzheimer’s disease. A widely available plasma-based AT(N) panel could support the triage of patients whose initial lab results and/or cognitive exams merit further evaluation of dementia. An AT(N) panel having sufficient analytical and clinical accuracy would allow for an initial, simple, blood-based test to provide evidence of AD pathology, which could then be confirmed with either a CSF test or PET scan. Such a panel could help identify AD patients much faster, shorten the overall AD patient diagnostic journey, enable earlier treatment interventions, and potentially reduce the enrollment duration and cost of clinical trials for AD therapeutics.

Although various analytes have been proposed for plasma-based assessments of brain pathophysiology, this study focuses on measurements using automated, high sensitivity immunoassay platforms for each respective AT(N) biomarker.^3,9–13,15,16^ Amyloid-beta 1-42 (Aβ42) and amyloid-beta 1-40 (Aβ40) are measured independently to determine the ratio of the protein isoforms (Aβ42/40) which has been shown to decrease with the onset of AD.^9,11,15,17–20^ In addition, measurements of microtubule-associated protein tau phosphorylated at threonine 181 (pTau181) and neurofilament light chain (NfL) were investigated as both have been shown to become elevated with progression of AD.^9,11,15,21–30^ Significant evidence for each of these analytes demonstrates their viability as respective biomarkers of the AT(N) framework. Notably, we confirm the concordance of a plasma Aβ42/40 ratio test, relative to amyloid PET status, in a well-characterized clinical cohort distinct from the initial investigation of clinical utility for this analytical platform.^17,31^

## 3. Methods

### 3.1. Assays Utilized

Measurements of Aβ40 and Aβ42 were performed using high sensitivity chemiluminescent enzyme immunoassays on a Sysmex^®^ HISCL-5000 instrument as previously reported.^17,31^ NfL and pTau181 measurements were performed using Roche Elecsys^®^ electrochemiluminescence immunoassays on a Roche cobas^®^ e801 instrument.^32^ Both instruments are considered high-throughput clinical autoanalyzers. The assays used have not yet received regulatory approval from the FDA although the Sysmex assays are CE marked and have received regulatory approval in Japan. All measurements were performed at the Center for Esoteric Testing of Labcorp (Burlington, NC). Analytical validation studies, based on guidance from the Clinical & Laboratory Standards Institute, were previously performed for each assay (Supplemental Table 1). All results described are a result of single measurements.

### 3.2. Clinical Specimens

To assess the clinical performance of the AT(N) panel, 200 specimens from subjects in the Australian Imaging, Biomarker & Lifestyle (AIBL) Study of Ageing were tested (Supplemental Table 2).^33,34^ The AIBL study was approved by various institutional ethics committees and all subjects provided written informed consent.^33,35^ For each subject, amyloid status (A±) was determined using PET imaging (where centiloid levels ≥ 26 were deemed positive) and specimens were collected intravenously using K3EDTA tubes (Sarstedt 01.1605.008) with prostaglandin E1 added (33 ng/mL of whole blood). Following centrifugation, plasma was removed, sub-aliquoted, and stored in deep frozen conditions (*i.e.* −80°C) until measurement.^33,34^

The neuropsychological information of each subject was reviewed following each visit using a clinical panel (using standardized clinical criteria) to determine the cognitive status of each subject: cognitively unimpaired (CU), mild cognitive impairment (MCI), Alzheimer’s disease dementia (ADD)^36^. The specimens utilized came from 75 CU/A-, 49 CU/A+, 26 MCI/A+, and 50 ADD/A+ subjects (Supplemental Table 3). As part of the neuropsychological information used to assess cognitive status, mini-mental state examinations (MMSE) were performed on each subject to provide a quantification of cognitive impairment.^37^ Note that 3 subjects (34, 47, and 75) provided specimens at 2 different time points and different clinical states (see Supplemental Table 3).

To further assess NfL levels with respect to age, two additional sets of samples were acquired. K2EDTA plasma specimens were collected from 41 subjects less than 50 years in age under an IRB approved protocol (WCG IRB protocol #520100174, Supplemental Table 4). MMSE scores were assumed to be negligible for these subjects. Additionally, 12 K2EDTA plasma specimens were purchased from ProteoGenex (Inglewood, CA); these were obtained from subjects that have been diagnosed with Alzheimer’s disease using one or more forms of brain structural or functional evaluation (Supplemental Table 5). Specimens were collected under ethical regulations and informed consent was obtained for each subject.^38^ Age and MMSE scores were provided for these subjects.

### 3.3. Statistical Analysis

Receiving operator characteristic (ROC) analysis was performed with respect to amyloid PET status to assess the clinical performance of each AT(N) biomarker. In addition, box-and-whisker plots (using the Tukey method for plotting the whiskers and outliers) were used to visually represent results while the Mann-Whitney U test was performed to determine statistical significance with the clinically defined groups (i.e. CU/A-, CU/A+, MCI/A+, ADD/A+). Statistical significance is represented using the following convention: * (p < 0.05), ** (p < 0.01), *** (p < 0.001), and **** (p < 0.0001).

Analysis with respect to amyloid status and MMSE scores was performed by subdividing scores into the following groups: negligible (26-30), mild (21-25), moderate (11-20), and severe (≤ 10) similar to previous reports^39^. Subjects outside of the AIBL cohort less than 50 years of age were assumed to be A− and have MMSE scores within the negligible region. Results from two A− subjects (subjects 155 and 126) were removed from MMSE analysis as they were classified as CU but had MMSE scores that fell into the mild region (Supplemental Table 2). In addition, results from moderate (11-20) and severe (≤ 10) scores were combined (M/S) from analysis utilizing results from only the AIBL cohort as only 3 subjects had results in the severe range (subjects 22, 46, 50).

## 4. Results

### 4.1. ROC Analysis

Assessment of results (Figure 1 and Supplemental Figure 1) indicated that each biomarker was able to demonstrate a high degree of statistical difference (p < 0.0001) between A− and A+ subjects. Decreasing Aβ42/40 values were observed with A+ subjects producing an area-under-the curve (AUC) of 0.941 with a maximum efficiency (*i.e.* accuracy) of 92.5% observed at a cutoff ratio of 0.102. Results for pTau181 produced an AUC of 0.847 with a maximum efficiency of 81.5% at a cutoff of 0.977 pg/mL. NfL results produced an AUC of 0.666 with a maximum efficiency of 68.5% at a cutoff of 3.21 pg/mL. Note that the assays utilized herein have not been standardized to produce comparable concentrations across different assays and platforms; hence the cutoffs proposed are not applicable to measurements made using other available assays (Supplemental Figure 2).

**Figure 1:**
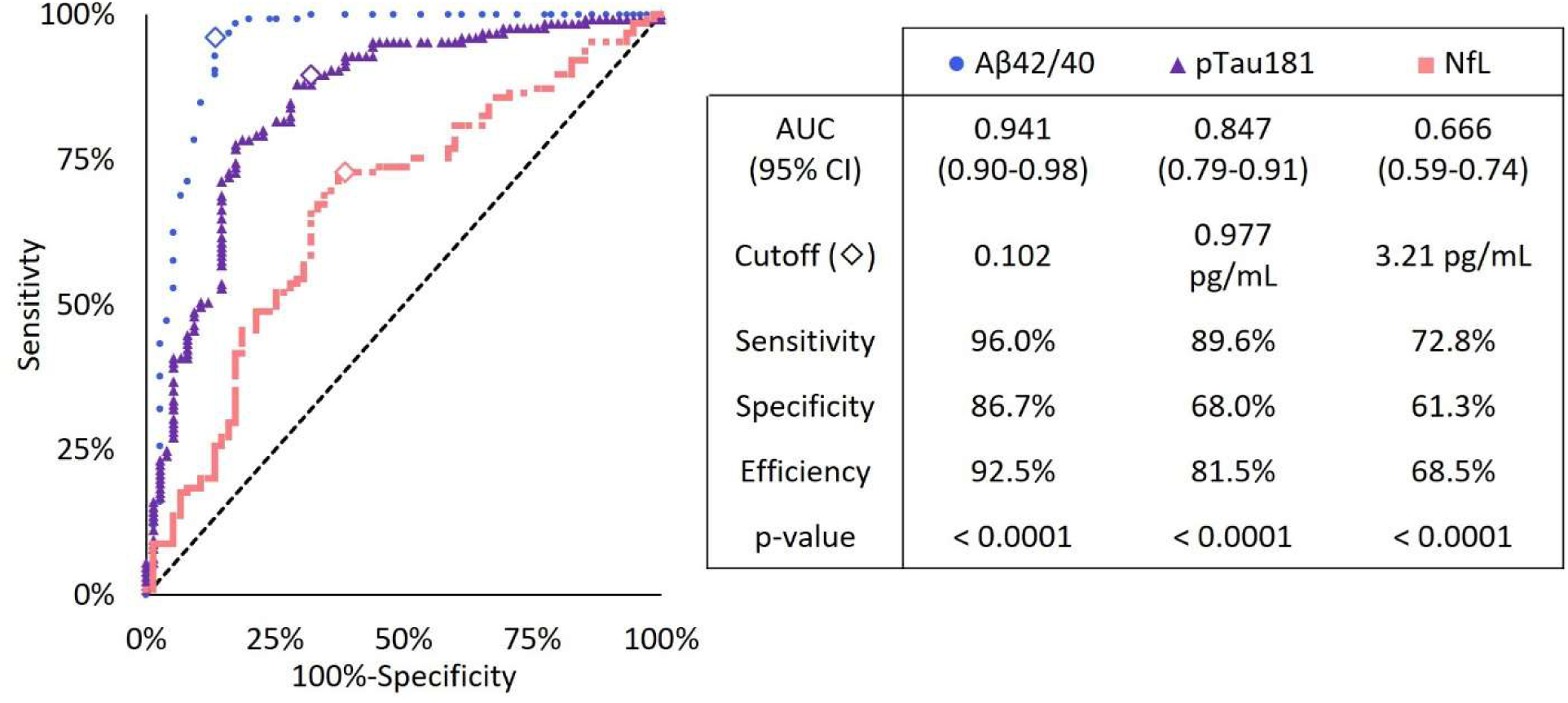
ROC plot of AT(N) biomarkers. Results are with respect to amyloid PET status and include the corresponding statistical results.

### 4.2. Amyloid and Cognitive Status Analysis

Investigation of Aβ42/40 results with respect to amyloid status as well as clinically defined cognitive status demonstrated a high significant difference (p < 0.0001) between A− and A+ subjects from each cognitive state (Figure 2A). There was also a difference observed between MCI/A+ subjects and CU/A+ as well as ADD/A+ subjects (p < 0.05) but not between CU/A+ and ADD/A+ subjects indicating that Aβ42/40 is likely not a good indicator of cognitive status. Results for pTau181 also demonstrated a high degree of significance (p < 0.0001) between A− and A+ subjects from each cognitive state (Figure 2B). However, pTau181 results from CU/A+ subjects were also found to be different than results from MCI/A+ subjects (p < 0.01) and ADD/A+ subjects (p < 0.0001). Results between the latter two groups were not statistically significant. NfL results for A− subjects as compared to the A+ subjects were statistically significant (p < 0.05) but the results between the different A+ groups were not (Figure 2C).

**Figure 2:**
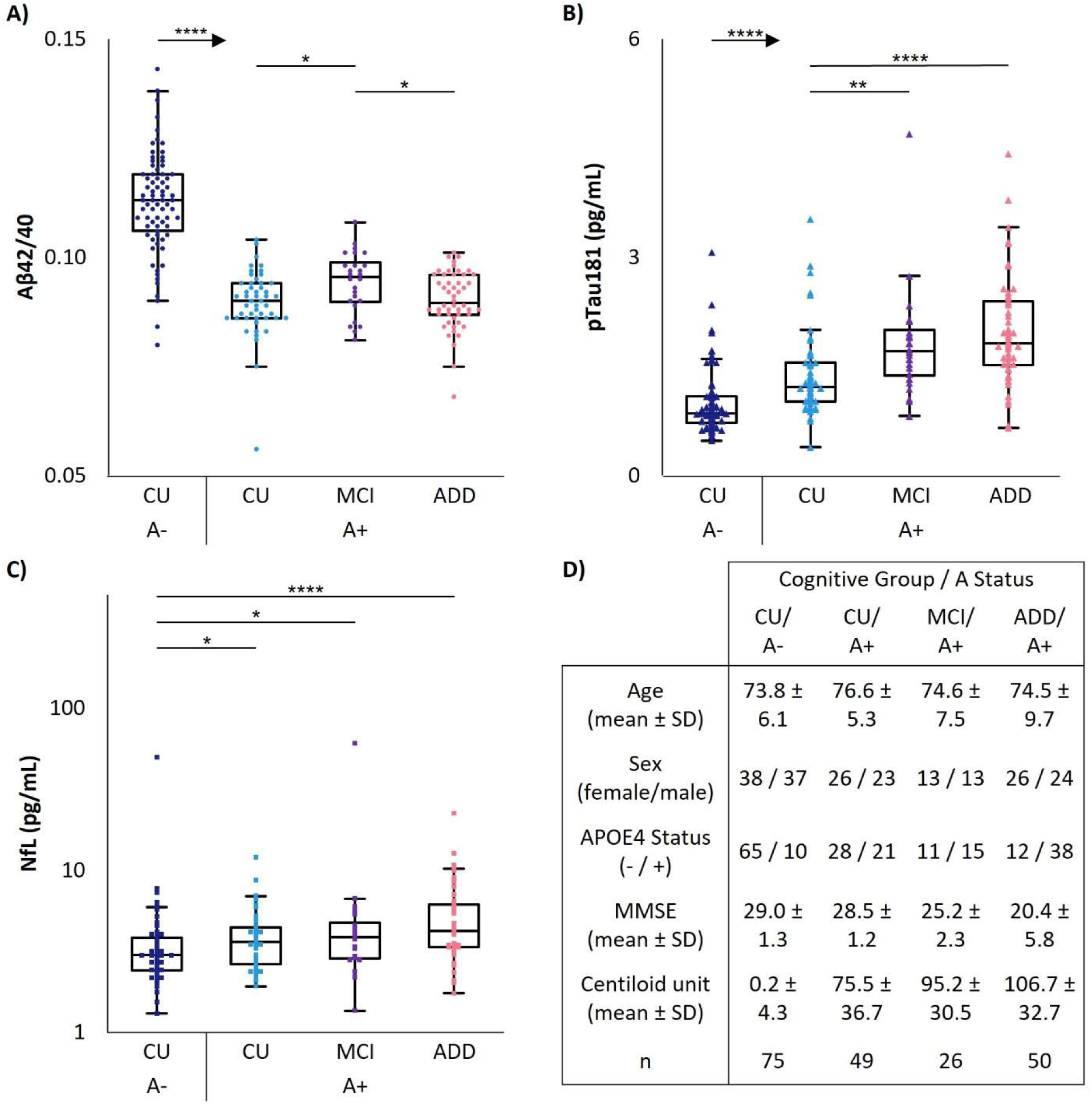
Results with respect to amyloid and clinically defined cognitive statuses. Results for A) Aβ42/40, B) pTau181, and C) NfL (note the logarithmic y-axis for NfL) are shown along with D) additional information for each of the groups analyzed.

### 4.3. Amyloid Status and MMSE Score Analysis

Similar to results observed with respect to amyloid and cognitive status, Aβ42/40 results differed between A− and A+ subjects with a high degree of significance (p < 0.0001) irrespective of MMSE scores (Figure 3A). However, results for A+ subjects from the negligible, mild, and moderate/severe groups were not statistically different. Results for pTau181 again demonstrated differences having a high degree of significance (p < 0.0001) between A− and A+ subjects from each MMSE range (Figure 23B). Results from negligible/A+ subjects were also found to be different than results from mild/A+ subjects (p < 0.05) and M/S/A+ subjects (p < 0.01). Results between the latter two groups were not statistically significant. NfL results, however, demonstrated statistical differences across all groups investigated (p < 0.05) with the exception A+ subjects having negligible and mild scores (Figure 3C). Similar results for each biomarker were observed with respect to centiloid values measured during PET analysis (Supplemental Figure 3).

**Figure 3:**
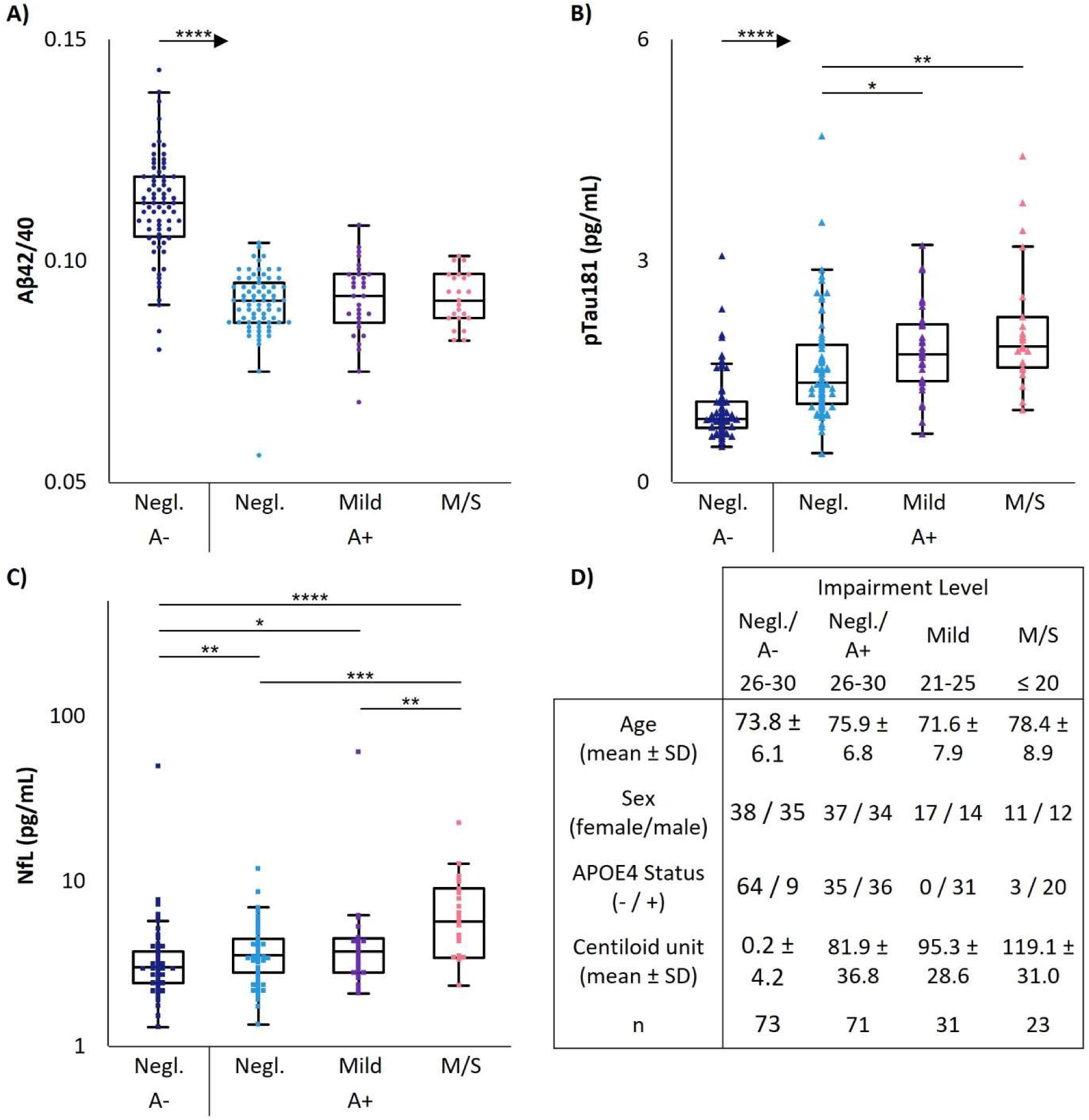
Results with respect to amyloid status and MMSE scores. Results for A) Aβ42/40, B) pTau181, and C) NfL (note the logarithmic y-axis for NfL) are shown along with D) additional information for each of the groups analyzed.

### 4.4. NfL Levels with Respect to Age and MMSE Scores

Expanding our analysis of NfL results to include additional younger subjects and more A+ subjects with severe MMSE scores (Figure 4), it is apparent that NfL levels are lower (p < 0.0001) in younger subjects (≤ 50 years) as compared to older subjects (> 50 years) regardless of amyloid status or MMSE scores. In addition, NfL levels from subjects with severe MMSE scores differed from subjects with moderate, mild, and negative MMSE scores with a high significant difference (p < 0.001). Older A− subjects with negligible MMSE scores also demonstrated statistically significant differences as compared to older A+ subjects with negligible and mild MMSE scores (p < 0.05). The significance increased when comparing older A− subjects with negligible MMSE scores to older A+ subjects with moderate MMSE scores (p < 0.0001). Older A+ subjects with negligible MMSE scores (p < 0.001) and older A+ subjects with mild MMSE scores (p < 0.01) also demonstrated a difference in NfL results as compared to older A+ subjects with moderate MMSE scores. Although a statistically significant difference was not observed between older A+ subjects with negligible and moderate MMSE scores, the results overall demonstrate an elevation of NfL with increased cognitive impairment.

**Figure 4:**
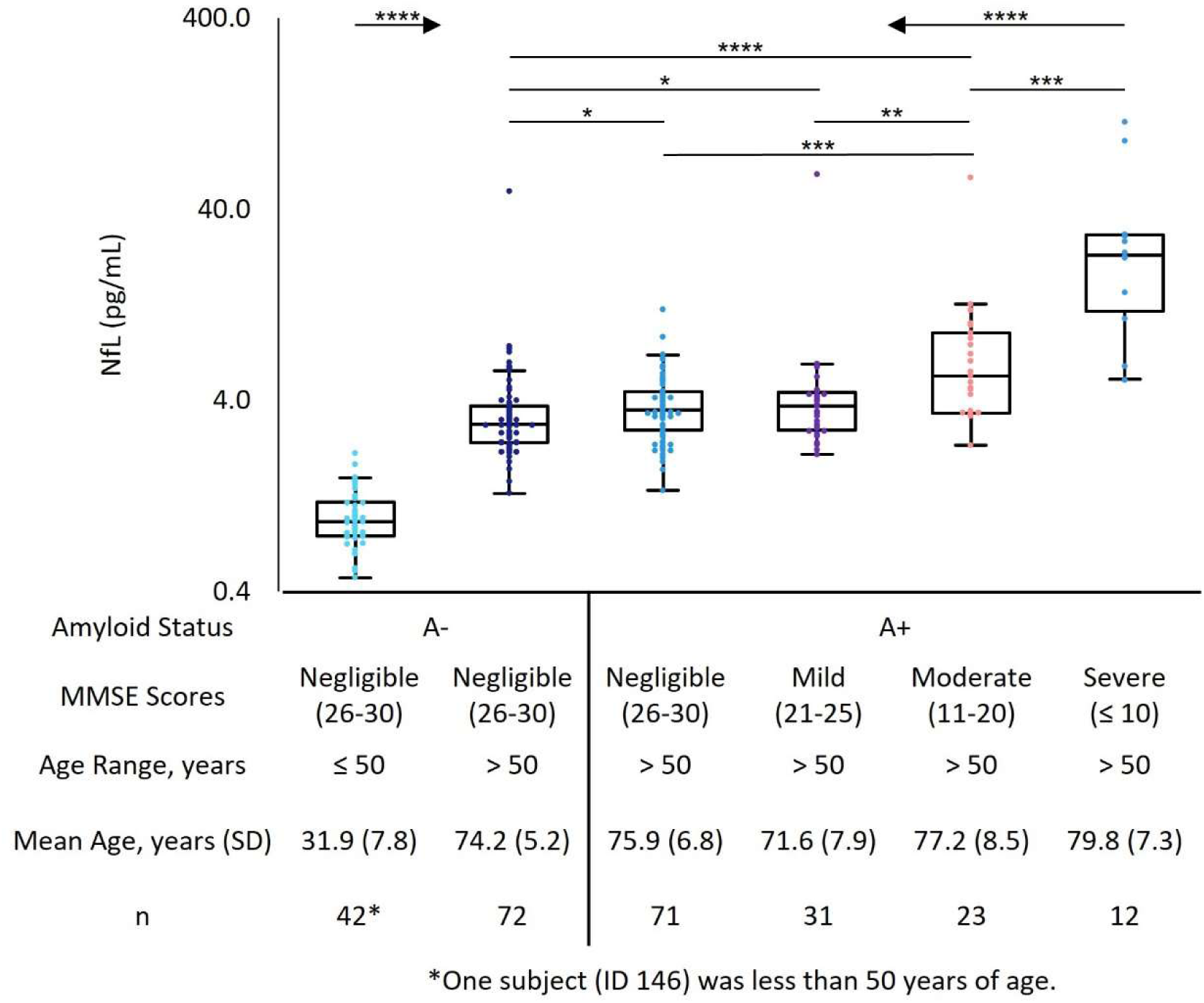
Expanded NfL Results. NfL results across multiple specimen groups with respect to amyloid status and MMSE scores (note the logarithmic y-axis for NfL). Additional information for each of the groups analyzed can be found below the graph.

## 5. Discussion

In order to assess relative performance of these assays, the observed AUC results were compared to those previously reported for ROC analyses performed with respect to amyloid status and irrespective of cognitive impairment. In addition, only results from standalone analysis of each biomarker (*i.e.* without incorporating additional measures or demographic information) were used. Unless otherwise noted, the reported AUCs were generated using different biomarker assays as well as different clinical cohorts where amyloid status was determined using a variety of techniques. In cases where multiple cohorts were utilized, only the comprehensive AUC (where available) is referenced herein.

Aβ42/40 results within this study yielded an AUC (0.941) greater than those reported for other Aβ42/40 measurement methods (0.64 to 0.89).^9,15,18,20,24,34,40–46^ It is interesting to note that the same Aβ42/40 measurement system and assays utilized herein produced similar AUC results (0.87-0.95) reported for different clinical cohorts with the same measurement system.^17,47^ In addition, the cutoff proposed in one of these studies matches the optimal Aβ42/40 ratio observed in the current study (0.102).^17^ Furthermore, the sensitivity (96.0%) and specificity (86.7%) observed herein for Aβ42/40 meets current recommendations for sensitivity (≥ 90.0%) and specificity (≥ 85.0%) for a triage test performed in a primary care setting.^48^ As Aβ42/40 is indicative of the early stages of AD progression (i.e. formation of amyloid plaques), it is not expected that it is an indicator of the progression of the disease from cognitively unimpaired to dementia nor indicative of lower MMSE scores.^49^ As such, Aβ42/40 levels appear to plateau following subjects becoming A+.

AUC results observed (0.847) for measurements of pTau181 are consistent with other reported AUC results (0.72 to 0.96).^9,10,23,24,34,40,43,46,47,50–53^ The AUC observed is also greater than the AUCs (0.77 to 0.80) previously reported using an assay from the same manufacturer but on a different instrument model (*i.e.* Roche cobas^®^ e601) with 2 different clinical cohorts.^9^ Despite the lower AUC values with respect to the AUC observed for Aβ42/40, pTau181 and other markers of tau pathology have been observed to be better predictors of cognitive and functional decline.^54–56^ In addition, the optimal cutoff observed with this cohort (0.977 pg/mL, Figure 1) is not statistically different than the cutoff established during initial validation studies (0.95 pg/mL, Supplemental Table 1)

Measurements of NfL on these AIBL samples demonstrated an AUC (0.666) consistent with those found in the literature (0.50 to 0.73).^9,40,46,47,52,57^ NfL results were also assessed with respect to pTau181 positivity, as defined using the pTau181 cutoff (0.977 pg/mL) determined during ROC analysis (Figure 1), as Tau positivity would indicate subjects that have progressed further along the biological framework of AD towards neurodegeneration. From this analysis, an AUC of 0.799 with p < 0.0001 was observed (Supplemental Figure 4). Elevation of both pTau181 and NfL levels was observed with worsening cognitive status or MMSE scores as has been previously reported.^26,30,58,59^ In addition, increases in NfL concentrations with respect to age has also been previously reported.^60^ These results coupled with the relatively low AUC of NfL (with respect to amyloid status) are further evidence that NfL may be an indicator of disease severity in AD.^12,21,26,29,51,61^

Although each biomarker investigated demonstrated the ability to distinguish between A− and A+ subjects, the AUCs were found to decrease with each AT(N) biomarker with respect to amyloid PET status. However, pTau181 and NfL correlated better with cognitive status than did the Aβ42/40 ratio (Figure 2 and Figure 3) suggesting that while the Aβ42/40 ratio may be central to differentiating AD from other diseases, pTau181 and NfL may provide some measure of disease severity.^12,21,26,27,29,51,61,62^ This is expected, as amyloid PET only indicates the presence of amyloid plaques within the brain and not the presence of other biological aspects of AD (*i.e.* tau pathologies and neurodegeneration). One shortcoming of this study is the lack of imaging endpoints for pTau181 and NfL (*e.g.* Tau PET or MRI) to make more appropriate correlative comparisons for each biomarker. However, age-dependent reference intervals can indicate if pathological levels exist especially for NfL where elevated levels are not AD specific (Supplemental Table 1).^16,63^

Although each of these biomarkers independently demonstrated clinical utility using specimens from subjects with known amyloid status, the combination of (quantitated) biomarkers with other factors such as demographics and the presence of the apolipoprotein E4 allele have been used to further improve the clinical accuracy of measurements.^9,14,52,64–67^ Additional biomarkers, such as tau protein phosphorylated at threonine 217 and glial fibrillary acidic protein, have also been reported to further characterize the underlying biology of AD as well as to assist in differentiating it from other forms of dementia.^5,14,16,56,56,67–70^

## 6. Conclusions

The AT(N) plasma panel is the first blood-based offering that analyzes three well-studied biomarkers to accurately detect biological evidence consistent with Alzheimer’s disease pathology. Furthermore, the sensitivity and specificity of the Aβ42/40 assay met current recommendations for triage testing. As such, the panel can be utilized to assess progression of subjects along the AT(N) framework when used within the context of a full clinical workup. The observed results indicate the advancements in and utility of blood-based biomarkers for assessing patients for Alzheimer’s disease. The availability of this panel serves as an important tool to assist in patient assessment and accelerate the path to diagnosis (and treatment) while the understanding of AD etiology and related pathophysiology continues to grow.

## Data Availability

All data discussed are contained within the manuscript or the supplemental information.

## Abbreviations

A±: amyloid status (positive or negative) based on PET imaging
Aβ: amyloid beta
Aβ40: amyloid beta consisting of amino acid residues 1-40
Aβ42: amyloid beta consisting of amino acid residues 1-42
Aβ42/40: the ratio of Aβ42 to Aβ40
AIBL: Australian Imaging, Biomarkers and Lifestyle (Study of Ageing)
AUC: area-under-the-curve
AD: Alzheimer’s disease
ADD: Alzheimer’s disease dementia
AT(N): amyloid, tau, neurodegeneration
CSF: cerebrospinal fluid
M/S: moderate or severe (with respect to MMSE scores)
MMSE: mini-mental state examination
Negl.: negligible (with respect to MMSE scores)
NfL: neurofilament light chain
PET: positron emission tomography
pTau181: microtubule-associated protein tau phosphorylated at threonine 181
ROC: receiver operator characteristic

## 8. Acknowledgements

The authors would like to thank Christopher Fowler and the AIBL Research Group for assistance in obtaining clinical specimens from the AIBL cohort, Roche for assistance with the NfL and pTau181 assays, and Sysmex for assistance with the Aβ40 and Aβ42 assays. In addition, the authors would like to thank Christos Petropolous of Monogram Biosciences (Labcorp) for his role in the acquisition and validation of AIBL samples as well as providing guidance in the validation and characterization of these assays. Furthermore, the authors would like to thank Ahmed Chenna of Monogram Biosciences for assistance with sample measurements using alternate instruments and assays as well as acquisition of samples. Finally, the authors would like to thank Amanda Suchanek, previously of Labcorp, for her initial work with the Sysmex^®^ instrument and assays.

