## Supplemental for "Clinical Assessment of a Plasma AT(N) Panel for Alzheimer’s Disease"

### Table 1: Analytical Validation Results Overview

| Study | Details | CLSI Guidance | Aβ42 | Aβ40 | pTau181 | NfL |
|---|---|---|---|---|---|---|
| Imprecision | 20 days, 2 runs/day, 2 replicates/run<br>Venous samples and control materials | EP05-A3 | CVs ≤ 5.3%* | CVs ≤ 5.7%* | CVs ≤ 4.6% | CVs ≤ 7.1% |
| Blood Matrix Interferences | Triglycerides | EP07-ED3 and EP37-ED1 | 1500 mg/dL | 1500 mg/dL | 1500 mg/dL | 1500 mg/dL |
| | Hemolysate | | 1000 mg/dL | 1000 mg/dL | 1500 mg/dL | 1500 mg/dL |
| | Total Protein | | 18.5 g/dL | 18.5 g/dL | 18.5 g/dL | 10.7 g/dL |
| | Bilirubin, conjugated | | 40 mg/dL | 40 mg/dL | 40 mg/dL | 40 mg/dL |
| | Bilirubin, unconjugated | | 40 mg/dL | 40 mg/dL | 40 mg/dL | 40 mg/dL |
| | Biotin | | 3510 ng/mL | 3510 ng/mL | 3510 ng/mL | 1170 ng/mL |
| Analytical Measurement Range | Lower limit of quantitation and dilutional linearity studies performed | EP17-A2 and EP06 | 3-2000 pg/mL | 25-2000 pg/mL | 0.3-10 pg/mL | 0.23-1000 pg/mL |
| Isothermal Stability | Room temperature (20 - 25 °C) | Not applicable | 4 hours | 4 hours | 14 days** | 14 days** |
| | Refrigerated (2 - 8 °C) | | 10 hours | 10 hours | 14 days** | 14 days** |
| | Frozen (< -10 °C) | | 24 hours | 24 hours | 14 days** | 14 days** |
| | Freeze/Thaw Cycles | | 1 cycle | 1 cycle | 3 cycles** | 3 cycles** |
| | Deep Frozen (< -70 °C) | | 100 days** | 100 days** | 63 days** | 199 days** |
| Method Repeatability | 20 samples measured in triplicate over a 3 day period<br>(1 measurement/day) | Not applicable | Mean CV = 3.1% | Mean CV = 3.1% | Mean CV = 2.5% | Mean CV = 3.7% |
| Reference Interval/Cutoff | More than 120 samples measured in each age group indicated | EP28-A3c | Not applicable** | Not applicable** | 0 to 55 years: <0.95 pg/mL<br>> 55 years: <0.97 pg/mL | 0 to 4 years: < 1.97 pg/mL<br>5 to 9 years: < 1.64 pg/mL<br>10 to 14 years: < 1.43 pg/mL<br>15 to 19 years: < 1.60 pg/mL<br>20 to 29 years: < 1.65 pg/mL<br>30 to 39 years: < 1.88 pg/mL<br>40 to 49 years: < 2.14 pg/mL<br>50 to 59 years: < 3.79 pg/mL<br>60 to 69 years: < 4.62 pg/mL<br>70 to 79 years: < 7.65 pg/mL<br>> 79 years: < 11.56 pg/mL |

*Corresponding CVs for Aβ42/40 were ≤ 6.5%, **Longest time point tested
**An Aβ42/40 ratio of > 0.102 implemented as a cutoff

**Table 2: List of AIBL Cohort Specimens including Demographics and AT(N) Results**

Note that in order to ensure de-identification of specimens, IDs have been changed from those provided by AIBL for the purposes of this manuscript.

| Specimen ID | Cognitive Status | Age Range (years) | MMSE Status | Centiloid Status | Amyloid Status | AB42/40 Result | pTau181 (pg/mL) | NfL (pg/mL) |
|---|---|---|---|---|---|---|---|---|
| 1 | ADD | 51-60 | Moderate | High | A+ | 0.084 | 2.11 | 3.43 |
| 2 | ADD | 51-60 | Mild | High | A+ | 0.075 | 2.14 | 2.85 |
| 3 | ADD | 71-80 | Mild | High | A+ | 0.088 | 1.89 | 5.98 |
| 4 | ADD | 81-90 | Moderate | Very High | A+ | 0.090 | 2.01 | 5.66 |
| 5 | ADD | 81-90 | Moderate | Very High | A+ | 0.101 | 1.30 | 6.41 |
| 6 | ADD | 71-80 | Moderate | Very High | A+ | 0.093 | 1.53 | 3.38 |
| 7 | ADD | 71-80 | Negligible | Very High | A+ | 0.092 | 1.47 | 3.40 |
| 8 | ADD | 71-80 | Mild | High | A+ | 0.094 | 1.02 | 2.78 |
| 9 | ADD | 51-60 | Negligible | High | A+ | 0.085 | 1.68 | 1.73 |
| 10 | ADD | 51-60 | Mild | High | A+ | 0.092 | 2.19 | 2.62 |
| 11 | ADD | 51-60 | Mild | High | A+ | 0.089 | 1.91 | 3.72 |
| 12 | ADD | 71-80 | Mild | Moderate | A+ | 0.099 | 1.37 | 2.65 |
| 13 | ADD | 51-60 | Mild | High | A+ | 0.068 | 2.44 | 2.98 |
| 14 | ADD | 51-60 | Moderate | High | A+ | 0.088 | 1.63 | 2.32 |
| 15 | ADD | 81-90 | Mild | Very High | A+ | 0.087 | 1.62 | 4.50 |
| 16 | ADD | 71-80 | Mild | Very High | A+ | 0.096 | 1.29 | 2.08 |
| 17 | ADD | 71-80 | Negligible | Very High | A+ | 0.098 | 2.57 | 3.40 |
| 18 | ADD | 51-60 | Negligible | High | A+ | 0.094 | 2.57 | 4.44 |
| 19 | ADD | 51-60 | Mild | High | A+ | 0.088 | 2.90 | 3.41 |
| 20 | ADD | 71-80 | Moderate | High | A+ | 0.084 | 1.78 | 3.34 |
| 21 | ADD | 71-80 | Negligible | Very High | A+ | 0.096 | 1.62 | 3.34 |
| 22 | ADD | 81-90 | Severe | Very High | A+ | 0.093 | 1.81 | 22.4 |
| 23 | ADD | 71-80 | Moderate | Very High | A+ | 0.100 | 1.78 | 3.51 |

| Specimen ID | Cognitive Status | Age Range (years) | MMSE Status | Centiloid Status | Amyloid Status | AB42/40 Result | pTau181 (pg/mL) | NfL (pg/mL) |
|---|---|---|---|---|---|---|---|---|
| 24 | ADD | 81-90 | Negligible | High | A+ | 0.089 | 1.53 | 3.47 |
| 25 | ADD | 71-80 | Moderate | High | A+ | 0.082 | 3.19 | 8.45 |
| 26 | ADD | 61-70 | Mild | Very High | A+ | 0.080 | 3.21 | 6.18 |
| 27 | ADD | 71-80 | Mild | Very High | A+ | 0.088 | 1.61 | 3.11 |
| 28 | ADD | 71-80 | Mild | High | A+ | 0.096 | 1.39 | 4.53 |
| 29 | ADD | 71-80 | Mild | Very High | A+ | 0.094 | 1.78 | 4.31 |
| 30 | ADD | 71-80 | Mild | High | A+ | 0.097 | 1.74 | 4.21 |
| 31 | ADD | 71-80 | Moderate | Very High | A+ | 0.087 | 1.97 | 7.00 |
| 32 | ADD | 71-80 | Mild | High | A+ | 0.083 | 1.05 | 2.94 |
| 33 | ADD | 71-80 | Negligible | High | A+ | 0.085 | 0.691 | 2.00 |
| 34B | ADD | 71-80 | Moderate | Very High | A+ | 0.091 | 0.977 | 3.43 |
| 35 | ADD | 71-80 | Negligible | Very High | A+ | 0.094 | 1.02 | 3.21 |
| 36 | ADD | 81-90 | Moderate | Very High | A+ | 0.097 | 3.41 | 7.82 |
| 37 | ADD | 81-90 | Moderate | Very High | A+ | 0.089 | 1.96 | 4.67 |
| 38 | ADD | 71-80 | Moderate | Very High | A+ | 0.087 | 2.24 | 4.56 |
| 39 | ADD | 81-90 | Moderate | High | A+ | 0.087 | 1.82 | 8.98 |
| 40 | ADD | 71-80 | Mild | High | A+ | 0.086 | 2.88 | 4.47 |
| 41 | ADD | 81-90 | Moderate | Very High | A+ | 0.100 | 1.55 | 3.37 |
| 42 | ADD | 71-80 | Moderate | High | A+ | 0.082 | 1.84 | 4.30 |
| 43 | ADD | 81-90 | Moderate | Very High | A+ | 0.096 | 1.09 | 9.89 |
| 44 | ADD | 71-80 | Negligible | Very High | A+ | 0.088 | 1.96 | 4.17 |
| 45 | ADD | 81-90 | Negligible | High | A+ | 0.084 | 2.35 | 6.09 |
| 46 | ADD | 81-90 | Severe | Very High | A+ | 0.088 | 1.46 | 6.04 |
| 47B | ADD | 81-90 | Moderate | Very High | A+ | 0.097 | 3.79 | 10.2 |
| 48 | ADD | 71-80 | Moderate | High | A+ | 0.097 | 1.93 | 5.38 |
| 49 | ADD | 81-90 | Moderate | High | A+ | 0.093 | 2.51 | 12.70 |
| 50 | ADD | 71-80 | Severe | Very High | A+ | 0.096 | 4.42 | 10.70 |

| Specimen ID | Cognitive Status | Age Range (years) | MMSE Status | Centiloid Status | Amyloid Status | AB42/40 Result | pTau181 (pg/mL) | NfL (pg/mL) |
|---|---|---|---|---|---|---|---|---|
| 51 | MCI | 61-70 | Mild | High | A+ | 0.095 | 1.35 | 3.99 |
| 52 | MCI | 61-70 | Negligible | Very High | A+ | 0.093 | 1.81 | 1.35 |
| 53 | MCI | 71-80 | Negligible | High | A+ | 0.098 | 1.33 | 3.26 |
| 54 | MCI | 61-70 | Mild | High | A+ | 0.083 | 1.69 | 2.19 |
| 55 | MCI | 71-80 | Negligible | Very High | A+ | 0.084 | 2.13 | 4.34 |
| 56 | MCI | 61-70 | Mild | Very High | A+ | 0.081 | 1.82 | 2.78 |
| 57 | MCI | 71-80 | Mild | Very High | A+ | 0.097 | 1.59 | 4.01 |
| 58 | MCI | 61-70 | Mild | Very High | A+ | 0.103 | 1.60 | 2.35 |
| 59 | MCI | 71-80 | Negligible | High | A+ | 0.101 | 1.27 | 4.33 |
| 60 | MCI | 61-70 | Mild | Very High | A+ | 0.102 | 2.12 | 4.31 |
| 61 | MCI | 91-100 | Negligible | Moderate | A+ | 0.089 | 1.48 | 5.91 |
| 62 | MCI | 81-90 | Mild | Very High | A+ | 0.101 | 2.38 | 3.50 |
| 63 | MCI | 61-70 | Negligible | Very High | A+ | 0.098 | 2.33 | 3.71 |
| 64 | MCI | 71-80 | Mild | High | A+ | 0.098 | 0.819 | 3.98 |
| 65 | MCI | 71-80 | Negligible | High | A+ | 0.096 | 1.66 | 5.31 |
| 66 | MCI | 71-80 | Mild | High | A+ | 0.090 | 1.25 | 3.32 |
| 67 | MCI | 71-80 | Negligible | High | A+ | 0.096 | 4.69 | 6.65 |
| 68 | MCI | 71-80 | Mild | Very High | A+ | 0.108 | 0.655 | 2.58 |
| 69 | MCI | 71-80 | Mild | High | A+ | 0.097 | 1.73 | 4.15 |
| 70 | MCI | 71-80 | Mild | High | A+ | 0.092 | 1.96 | 60.7 |
| 47A | MCI | 81-90 | Mild | Very High | A+ | 0.095 | 2.47 | 6.13 |
| 71 | MCI | 71-80 | Negligible | Very High | A+ | 0.101 | 2.74 | 5.54 |
| 72 | MCI | 81-90 | Negligible | Very High | A+ | 0.084 | 1.90 | 3.36 |
| 73 | MCI | 71-80 | Mild | High | A+ | 0.085 | 1.53 | 5.29 |
| 74 | MCI | 71-80 | Negligible | Very High | A+ | 0.091 | 1.19 | 2.17 |
| 75B | CU | 71-80 | Negligible | Moderate | A+ | 0.103 | 0.752 | 2.59 |
| 76 | CU | 71-80 | Negligible | High | A+ | 0.098 | 1.67 | 4.08 |

| Specimen ID | Cognitive Status | Age Range (years) | MMSE Status | Centiloid Status | Amyloid Status | AB42/40 Result | pTau181 (pg/mL) | NfL (pg/mL) |
|---|---|---|---|---|---|---|---|---|
| 77 | CU | 71-80 | Negligible | Very High | A+ | 0.098 | 1.26 | 2.91 |
| 78 | CU | 71-80 | Negligible | High | A+ | 0.096 | 1.09 | 2.79 |
| 79 | CU | 91-100 | Negligible | Very High | A+ | 0.089 | 2.88 | 6.06 |
| 80 | CU | 71-80 | Negligible | Moderate | A+ | 0.086 | 1.14 | 4.82 |
| 81 | CU | 71-80 | Negligible | High | A+ | 0.093 | 1.55 | 1.91 |
| 82 | CU | 71-80 | Negligible | Moderate | A+ | 0.094 | 1.89 | 4.12 |
| 83 | CU | 71-80 | Negligible | High | A+ | 0.075 | 1.42 | 3.38 |
| 84 | CU | 81-90 | Negligible | Moderate | A+ | 0.096 | 1.65 | 5.03 |
| 85 | CU | 71-80 | Negligible | Very High | A+ | 0.091 | 0.95 | 2.07 |
| 34A | CU | 71-80 | Negligible | Moderate | A+ | 0.086 | 1.37 | 3.43 |
| 86 | CU | 81-90 | Negligible | Moderate | A+ | 0.083 | 1.70 | 4.46 |
| 87 | CU | 71-80 | Negligible | High | A+ | 0.104 | 0.880 | 3.97 |
| 88 | CU | 61-70 | Negligible | High | A+ | 0.087 | 2.79 | 3.96 |
| 89 | CU | 71-80 | Negligible | Moderate | A+ | 0.094 | 1.02 | 6.56 |
| 90 | CU | 81-90 | Negligible | Moderate | A+ | 0.094 | 1.51 | 4.09 |
| 91 | CU | 71-80 | Negligible | High | A+ | 0.092 | 1.06 | 2.19 |
| 92 | CU | 81-90 | Negligible | High | A+ | 0.089 | 1.20 | 4.31 |
| 93 | CU | 61-70 | Negligible | Moderate | A+ | 0.094 | 0.931 | 2.78 |
| 94 | CU | 81-90 | Negligible | High | A+ | 0.091 | 2.51 | 5.90 |
| 95 | CU | 71-80 | Negligible | High | A+ | 0.086 | 1.35 | 3.55 |
| 96 | CU | 71-80 | Negligible | High | A+ | 0.097 | 1.21 | 4.12 |
| 97 | CU | 81-90 | Negligible | High | A+ | 0.087 | 1.53 | 8.61 |
| 98 | CU | 71-80 | Negligible | Very High | A+ | 0.090 | 0.932 | 2.40 |
| 99 | CU | 71-80 | Negligible | Very High | A+ | 0.095 | 1.20 | 3.76 |
| 100 | CU | 71-80 | Negligible | High | A+ | 0.086 | 1.05 | 2.35 |
| 101 | CU | 71-80 | Negligible | High | A+ | 0.085 | 1.22 | 3.46 |
| 102 | CU | 71-80 | Negligible | Very High | A+ | 0.086 | 2.48 | 6.91 |

| Specimen ID | Cognitive Status | Age Range (years) | MMSE Status | Centiloid Status | Amyloid Status | AB42/40 Result | pTau181 (pg/mL) | NfL (pg/mL) |
|---|---|---|---|---|---|---|---|---|
| 103 | CU | 81-90 | Negligible | High | A+ | 0.091 | 0.789 | 2.29 |
| 104 | CU | 71-80 | Negligible | Moderate | A+ | 0.083 | 1.03 | 2.65 |
| 105 | CU | 81-90 | Negligible | High | A+ | 0.092 | 0.907 | 2.43 |
| 106 | CU | 81-90 | Negligible | Very High | A+ | 0.086 | 3.52 | 11.9 |
| 107 | CU | 71-80 | Negligible | Very High | A+ | 0.087 | 2.00 | 3.81 |
| 108 | CU | 81-90 | Negligible | High | A+ | 0.093 | 1.16 | 5.15 |
| 109 | CU | 71-80 | Negligible | High | A+ | 0.088 | 1.30 | 2.98 |
| 110 | CU | 71-80 | Negligible | High | A+ | 0.100 | 1.33 | 3.46 |
| 111 | CU | 71-80 | Negligible | High | A+ | 0.090 | 1.54 | 4.18 |
| 112 | CU | 71-80 | Negligible | High | A+ | 0.091 | 0.392 | 2.35 |
| 113 | CU | 71-80 | Negligible | High | A+ | 0.056 | 1.55 | 4.37 |
| 114 | CU | 71-80 | Negligible | High | A+ | 0.086 | 1.10 | 3.28 |
| 115 | CU | 61-70 | Negligible | Moderate | A+ | 0.082 | 0.935 | 2.17 |
| 116 | CU | 71-80 | Negligible | Moderate | A+ | 0.092 | 1.27 | 2.58 |
| 117 | CU | 71-80 | Negligible | Moderate | A+ | 0.081 | 1.02 | 2.27 |
| 118 | CU | 71-80 | Negligible | High | A+ | 0.083 | 1.20 | 3.59 |
| 119 | CU | 71-80 | Negligible | Very High | A+ | 0.089 | 1.86 | 6.61 |
| 120 | CU | 81-90 | Negligible | Very High | A+ | 0.091 | 1.27 | 3.63 |
| 121 | CU | 71-80 | Negligible | Very High | A+ | 0.097 | 0.992 | 5.18 |
| 122 | CU | 71-80 | Negligible | Moderate | A+ | 0.087 | 0.907 | 3.40 |
| 123 | CU | 81-90 | Negligible | High | A+ | 0.090 | 1.92 | 3.28 |
| 124 | CU | 71-80 | Negligible | Negative | A- | 0.121 | 0.689 | 3.09 |
| 75A | CU | 71-80 | Negligible | Negative | A- | 0.113 | 1.08 | 2.14 |
| 125 | CU | 61-70 | Negligible | Negative | A- | 0.120 | 0.751 | 2.41 |
| 126 | CU | 71-80 | Mild | Negative | A- | 0.108 | 1.07 | 3.82 |
| 127 | CU | 71-80 | Negligible | Negative | A- | 0.102 | 0.659 | 2.40 |
| 128 | CU | 71-80 | Negligible | Negative | A- | 0.097 | 1.16 | 2.97 |

| Specimen ID | Cognitive Status | Age Range (years) | MMSE Status | Centiloid Status | Amyloid Status | AB42/40 Result | pTau181 (pg/mL) | NfL (pg/mL) |
|---|---|---|---|---|---|---|---|---|
| 129 | CU | 71-80 | Negligible | Negative | A- | 0.122 | 1.02 | 3.63 |
| 130 | CU | 81-90 | Negligible | Negative | A- | 0.098 | 1.26 | 5.86 |
| 131 | CU | 71-80 | Negligible | Negative | A- | 0.103 | 0.863 | 3.32 |
| 132 | CU | 61-70 | Negligible | Negative | A- | 0.126 | 0.761 | 5.13 |
| 133 | CU | 61-70 | Negligible | Negative | A- | 0.090 | 1.72 | 1.90 |
| 134 | CU | 81-90 | Negligible | Negative | A- | 0.105 | 1.58 | 4.14 |
| 135 | CU | 71-80 | Negligible | Negative | A- | 0.084 | 2.00 | 4.00 |
| 136 | CU | 71-80 | Negligible | Negative | A- | 0.132 | 0.914 | 2.35 |
| 137 | CU | 61-70 | Negligible | Negative | A- | 0.121 | 0.799 | 2.16 |
| 138 | CU | 61-70 | Negligible | Negative | A- | 0.136 | 0.911 | 3.06 |
| 139 | CU | 81-90 | Negligible | Negative | A- | 0.112 | 0.661 | 3.35 |
| 140 | CU | 71-80 | Negligible | Negative | A- | 0.111 | 0.806 | 4.52 |
| 141 | CU | 71-80 | Negligible | Negative | A- | 0.111 | 0.740 | 2.15 |
| 142 | CU | 61-70 | Negligible | Negative | A- | 0.126 | 0.862 | 2.33 |
| 143 | CU | 71-80 | Negligible | Negative | A- | 0.138 | 0.948 | 4.00 |
| 144 | CU | 71-80 | Negligible | Negative | A- | 0.143 | 1.15 | 3.81 |
| 145 | CU | 71-80 | Negligible | Negative | A- | 0.116 | 0.820 | 2.75 |
| 146 | CU | 41-50 | Negligible | Negative | A- | 0.115 | 0.625 | 1.30 |
| 147 | CU | 71-80 | Negligible | Negative | A- | 0.114 | 0.850 | 2.95 |
| 148 | CU | 81-90 | Negligible | Negative | A- | 0.109 | 0.794 | 2.64 |
| 149 | CU | 61-70 | Negligible | Negative | A- | 0.123 | 1.00 | 2.69 |
| 150 | CU | 71-80 | Negligible | Negative | A- | 0.118 | 0.669 | 2.95 |
| 151 | CU | 71-80 | Negligible | Negative | A- | 0.119 | 0.953 | 2.58 |
| 152 | CU | 71-80 | Negligible | Negative | A- | 0.123 | 0.831 | 3.18 |
| 153 | CU | 71-80 | Negligible | Negative | A- | 0.107 | 0.917 | 2.73 |
| 154 | CU | 71-80 | Negligible | Negative | A- | 0.119 | 0.865 | 3.11 |
| 155 | CU | 71-80 | Mild | Negative | A- | 0.118 | 0.616 | 2.39 |

| Specimen ID | Cognitive Status | Age Range (years) | MMSE Status | Centiloid Status | Amyloid Status | AB42/40 Result | pTau181 (pg/mL) | NfL (pg/mL) |
|---|---|---|---|---|---|---|---|---|
| 156 | CU | 71-80 | Negligible | Negative | A- | 0.116 | 0.935 | 2.90 |
| 157 | CU | 71-80 | Negligible | Negative | A- | 0.124 | 0.956 | 3.25 |
| 158 | CU | 71-80 | Negligible | Negative | A- | 0.096 | 0.748 | 2.16 |
| 159 | CU | 71-80 | Negligible | Negative | A- | 0.115 | 0.756 | 3.57 |
| 160 | CU | 71-80 | Negligible | Negative | A- | 0.113 | 2.35 | 5.72 |
| 161 | CU | 71-80 | Negligible | Negative | A- | 0.129 | 0.838 | 2.95 |
| 162 | CU | 71-80 | Negligible | Negative | A- | 0.095 | 1.09 | 2.56 |
| 163 | CU | 61-70 | Negligible | Negative | A- | 0.105 | 0.556 | 2.41 |
| 164 | CU | 71-80 | Negligible | Negative | A- | 0.111 | 1.24 | 7.21 |
| 165 | CU | 71-80 | Negligible | Negative | A- | 0.112 | 0.665 | 2.38 |
| 166 | CU | 71-80 | Negligible | Negative | A- | 0.114 | 0.627 | 2.15 |
| 167 | CU | 71-80 | Negligible | Negative | A- | 0.117 | 0.626 | 2.03 |
| 168 | CU | 71-80 | Negligible | Negative | A- | 0.119 | 0.750 | 3.28 |
| 169 | CU | 71-80 | Negligible | Negative | A- | 0.124 | 1.60 | 6.02 |
| 170 | CU | 71-80 | Negligible | Negative | A- | 0.119 | 0.853 | 3.66 |
| 171 | CU | 71-80 | Negligible | Negative | A- | 0.114 | 0.647 | 3.94 |
| 172 | CU | 71-80 | Negligible | Negative | A- | 0.080 | 1.14 | 3.53 |
| 173 | CU | 71-80 | Negligible | Negative | A- | 0.104 | 0.653 | 2.34 |
| 174 | CU | 71-80 | Negligible | Negative | A- | 0.109 | 0.838 | 2.17 |
| 175 | CU | 71-80 | Negligible | Negative | A- | 0.107 | 0.845 | 3.00 |
| 176 | CU | 71-80 | Negligible | Negative | A- | 0.106 | 0.762 | 3.37 |
| 177 | CU | 71-80 | Negligible | Negative | A- | 0.104 | 1.65 | 3.46 |
| 178 | CU | 61-70 | Negligible | Negative | A- | 0.112 | 0.481 | 2.25 |
| 179 | CU | 51-60 | Negligible | Negative | A- | 0.111 | 0.596 | 1.52 |
| 180 | CU | 71-80 | Negligible | Negative | A- | 0.122 | 3.07 | 49.70 |
| 181 | CU | 61-70 | Negligible | Negative | A- | 0.118 | 0.726 | 2.40 |
| 182 | CU | 71-80 | Negligible | Negative | A- | 0.114 | 0.943 | 3.19 |

| Specimen ID | Cognitive Status | Age Range (years) | MMSE Status | Centiloid Status | Amyloid Status | AB42/40 Result | pTau181 (pg/mL) | NfL (pg/mL) |
|---|---|---|---|---|---|---|---|---|
| 183 | CU | 71-80 | Negligible | Negative | A- | 0.105 | 1.09 | 3.15 |
| 184 | CU | 71-80 | Negligible | Negative | A- | 0.108 | 0.590 | 2.21 |
| 185 | CU | 71-80 | Negligible | Negative | A- | 0.116 | 0.679 | 2.99 |
| 186 | CU | 81-90 | Negligible | Negative | A- | 0.094 | 1.55 | 5.96 |
| 187 | CU | 71-80 | Negligible | Negative | A- | 0.107 | 0.544 | 1.75 |
| 188 | CU | 71-80 | Negligible | Negative | A- | 0.091 | 1.11 | 3.15 |
| 189 | CU | 81-90 | Negligible | Negative | A- | 0.117 | 0.860 | 4.72 |
| 190 | CU | 71-80 | Negligible | Negative | A- | 0.109 | 0.909 | 2.69 |
| 191 | CU | 71-80 | Negligible | Negative | A- | 0.098 | 0.822 | 2.98 |
| 192 | CU | 71-80 | Negligible | Negative | A- | 0.109 | 1.96 | 7.68 |
| 193 | CU | 81-90 | Negligible | Negative | A- | 0.113 | 1.55 | 5.92 |
| 194 | CU | 71-80 | Negligible | Negative | A- | 0.102 | 1.01 | 2.95 |
| 195 | CU | 71-80 | Negligible | Negative | A- | 0.113 | 0.850 | 6.28 |
| 196 | CU | 71-80 | Negligible | Negative | A- | 0.127 | 0.718 | 2.52 |
| 197 | CU | 71-80 | Negligible | Negative | A- | 0.109 | 1.71 | 7.42 |

**Table 3: Overview of AIBL Cohort Specimens**

| Amyloid Status: | Negative | Positive | | | |
| --- | --- | --- | --- | --- | --- |
| **Cognitive Status:** | **CU** | **All** | **CU** | **MCI** | **ADD** |
| n: | 75 | 125 | 49 | 26 | 50 |
| Age (mean ± SD): | 73.8 ± 6.1 | 75.3 ± 7.8 | 76.6 ± 5.3 | 74.6 ± 7.5 | 74.5 ± 9.7 |
| Sex (female/male): | 38 / 37 | 65 / 60 | 26 / 23 | 13 / 13 | 26 / 24 |
| APOE4 Status (- / +): | 65 / 10 | 74 / 51 | 28 / 21 | 11 / 15 | 12 / 38 |
| Centiloid units (mean ± SD): | 0.2 ± 4.3 | 92.1 ± 36.4 | 28.5 ± 1.2 | 25.2 ± 2.3 | 20.4 ± 5.8 |
| MMSE Score (mean ± SD): | 29.0 ± 1.3 | 24.6 ± 5.3 | 75.5 ± 36.7 | 95.2 ± 30.5 | 106.7 ± 32.7 |
| [Aβ42/40]* | 0.113 (0.090 - 0.138) | 0.091 (0.075 - 0.108) | 0.090 (0.075 - 0.104) | 0.096 (0.081 - 0.108) | 0.090 (0.075 - 0.101) |
| [pTau181] pg/mL* | 0.853 (0.481 - 1.60) | 1.59 (0.392 - 2.90) | 1.22 (0.392 - 2.00) | 1.71 (0.655 - 2.74) | 1.82 (0.691 - 3.21) |
| [NfL] pg/mL* | 2.99 (1.30 - 5.92) | 3.76 (1.35 - 8.45) | 3.29 (1.91 - 6.91) | 3.99 (1.35 - 6.65) | 4.19 (1.73 - 9.89) |

*Values represent the median observed with the interquartile range as determined using the Tukey method in parentheses

**Table 4: Additional Specimens Utilized from Cognitively Unimpaired Subjects**

These subjects were assumed to be A- and have negligible MMSE scores.

| Specimen ID | Age Range (years) | NfL (pg/mL) |
|---|---|---|
| **LC01** | 31-40 | 0.818 |
| **LC02** | 31-40 | 1.12 |
| **LC03** | 31-40 | 1.45 |
| **LC04** | 31-40 | 1.22 |
| **LC05** | 21-30 | 0.888 |
| **LC06** | 41-50 | 1.07 |
| **LC07** | 41-50 | 2.1 |
| **LC08** | 41-50 | 1.56 |
| **LC09** | 21-30 | 0.873 |
| **LC10** | 21-30 | 1.03 |
| **LC11** | 21-30 | 1.17 |
| **LC12** | 21-30 | 1.50 |
| **LC13** | 21-30 | 1.27 |
| **LC14** | 21-30 | 0.510 |
| **LC15** | 41-50 | 1.84 |
| **LC16** | 21-30 | 0.471 |
| **LC17** | 21-30 | 0.929 |
| **LC18** | 41-50 | 0.707 |
| **LC19** | 31-40 | 1.00 |
| **LC20** | 31-40 | 0.797 |
| **LC21** | 31-40 | 0.848 |

| Specimen ID | Age Range (years) | NfL (pg/mL) |
|---|---|---|
| **LC22** | 21-30 | 0.963 |
| **LC23** | 41-50 | 1.17 |
| **LC24** | 21-30 | 0.946 |
| **LC25** | 21-30 | 0.709 |
| **LC26** | 21-30 | 0.784 |
| **LC27** | 21-30 | 0.761 |
| **LC28** | 31-40 | 0.81 |
| **LC29** | 31-40 | 0.956 |
| **LC30** | 21-30 | 0.527 |
| **LC31** | 21-30 | 0.769 |
| **LC32** | 21-30 | 0.625 |
| **LC33** | 41-50 | 1.38 |
| **LC34** | 21-30 | 0.915 |
| **LC35** | 21-30 | 0.774 |
| **LC36** | 21-30 | 0.783 |
| **LC37** | 21-30 | 1.57 |
| **LC38** | 21-30 | 0.922 |
| **LC39** | 41-50 | 0.981 |
| **LC40** | 21-30 | 0.808 |
| **LC41** | 21-30 | 0.658 |

**Table 5: Additional AD Specimens Utilized**

Specimens acquired from ProteoGenex (Inglewood, CA) were obtained from subjects that have been diagnosed with Alzheimer's disease using one or more forms of neurological evaluation. MMSE scores were also provided for these subjects.

| Specimen ID | Age Range (years) | Amyloid Status | Diagnostic Tests | MMSE Status | NfL (pg/mL) |
|---|---|---|---|---|---|
| **AG196S** | 81-90 | A+ | CT scan | Severe | 29.1 |
| **AG349S** | 71-80 | A+ | MRI, EEG | Moderate | 4.97 |
| **AG143S** | 81-90 | A+ | CT scan, EEG | Severe | 22.6 |
| **AG144S** | 71-80 | A+ | CT scan | Severe | 23.6 |
| **AG181S** | 61-70 | A+ | CT scan, EEG | Severe | 27.1 |
| **AG323S** | 71-80 | A+ | CT scan | Severe | 14.6 |
| **AG219S** | 81-90 | A+ | CT scan, EEG | Severe | 29.4 |
| **AG212S** | 81-90 | A+ | CT scan, EEG | Severe | 5.15 |
| **AG204S** | 71-80 | A+ | CT scan, EEG | Severe | 91 |
| **AG194S** | 71-80 | A+ | MRI | Moderate | 58.6 |
| **AG445S** | 71-80 | A+ | MRI | Moderate | 11.8 |
| **AG337S** | 71-80 | A+ | CT scan | Severe | 115 |

**Figure 1: AT(N) Results with Respect to Amyloid Status**

A) Aβ42/40, B) pTau181, and C) NfL (note the logarithmic y-axis for NfL) results of AIBL groups defined by amyloid status.

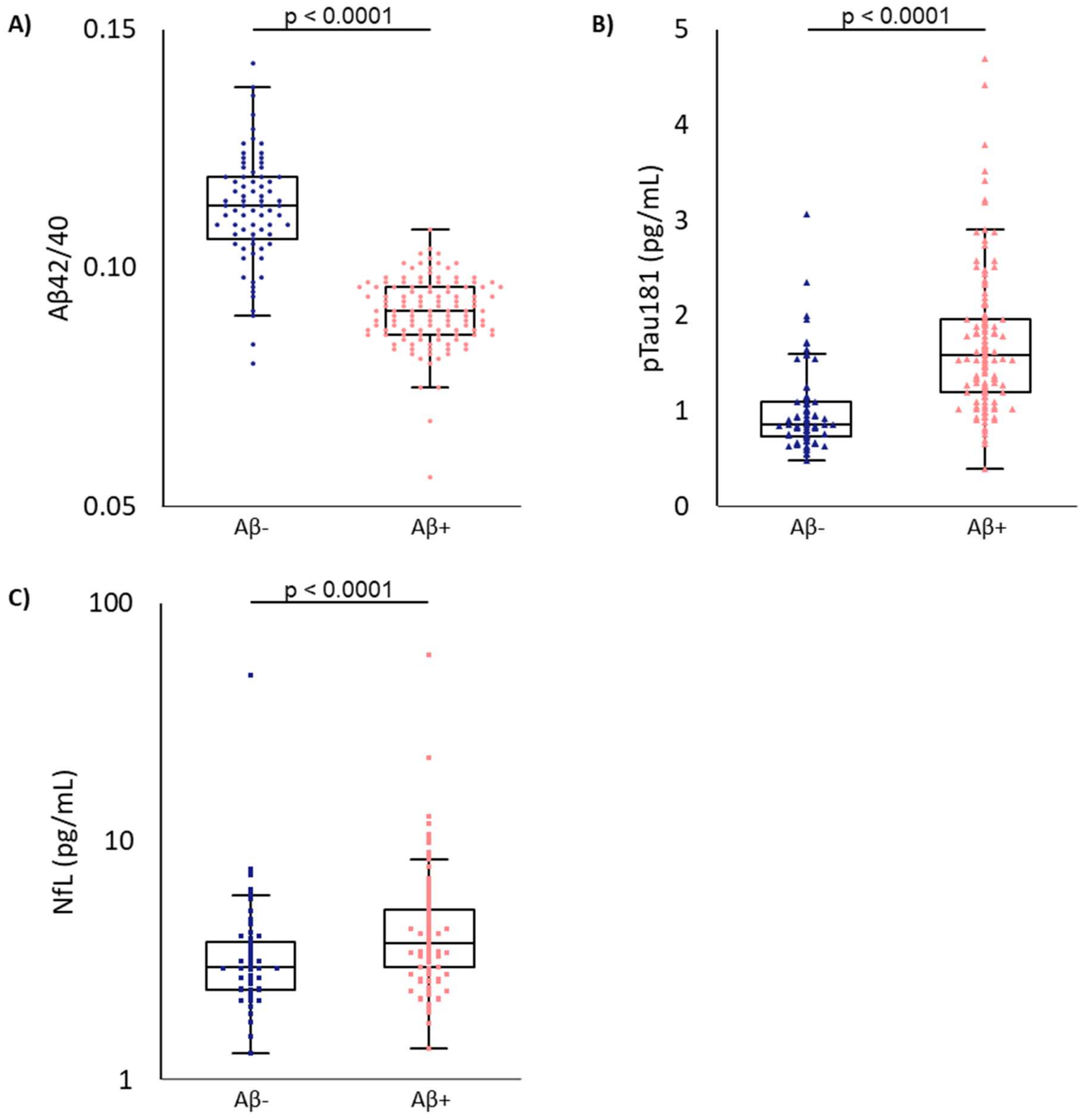

**Figure 2: Correlative Results of AIBL Specimens**

The AIBL cohort specimens used were also evaluated using commercially available assays for the same biomarkers on another platform. Results demonstrate correlations (R) > 0.81 for all individual measurements (i.e. excluding Aβ42/40 ratio).

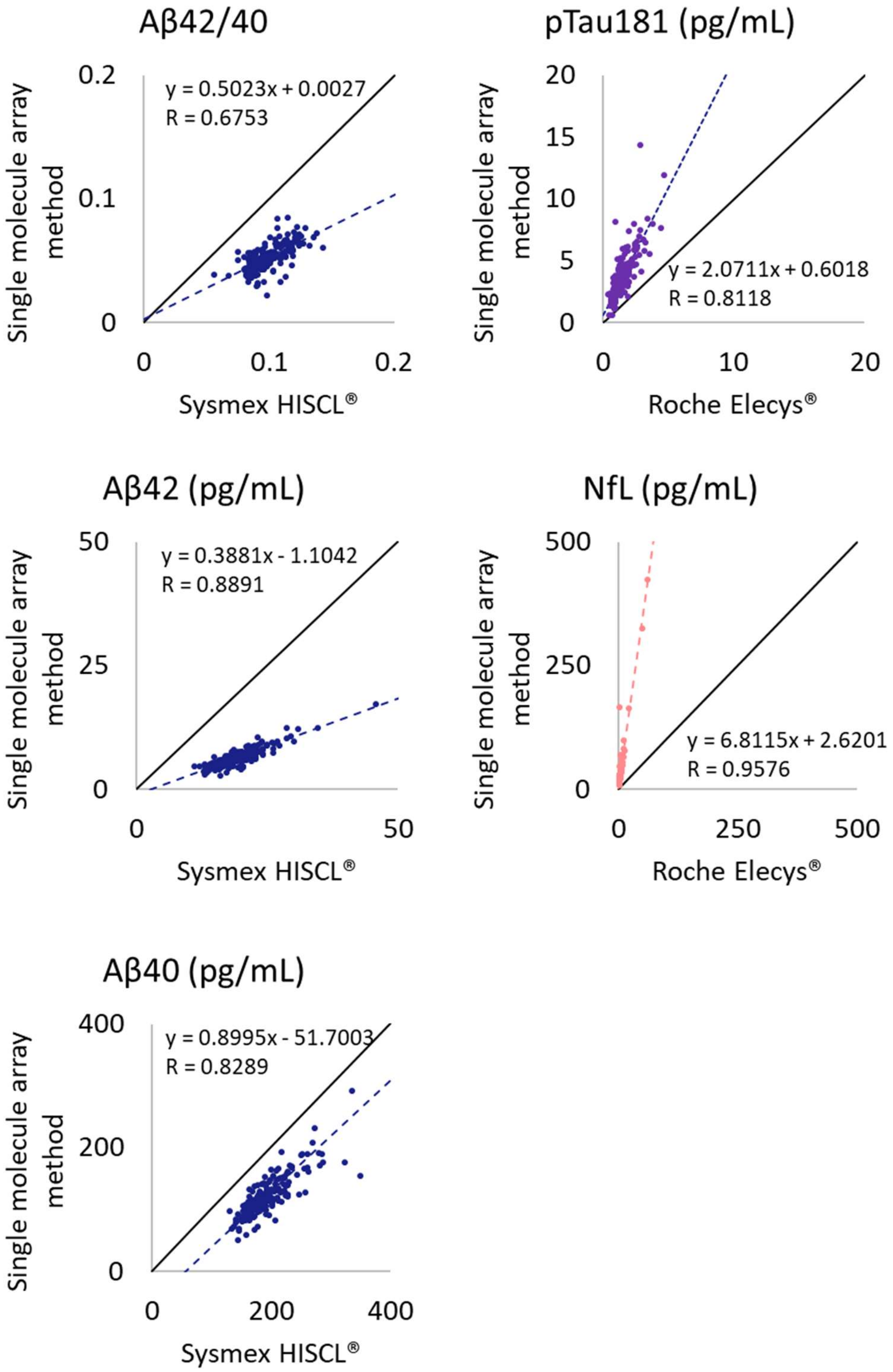

**Figure 3: Centiloid Analysis by Group**

Centiloid values obtained during PET imaging were used to assess biomarker results where centiloid levels (CL) were divided into standardized groups as follows: negative (< 15 CL), moderate (26-50 CL), high (51-100 CL), and very high (>100 CL). A) Aβ42/40 results were able to distinguish between negative and moderate to very high centiloid levels with a high degree of significance (p<0.0001). However, the significance between the results from A+ subjects was either not significant or produced a low degree of significance (p<0.05). B) Increases in centiloid values lead to an increase in pTau181 levels for all centiloid groups except for high and very high centiloid values where a significant difference was not observed. C) NfL levels were higher for the high and very high centiloid groups as compared to the negative group (p < 0.01), however, NfL levels across the A+ subjects were not significantly different. D) Additional information for each of the groups analyzed.

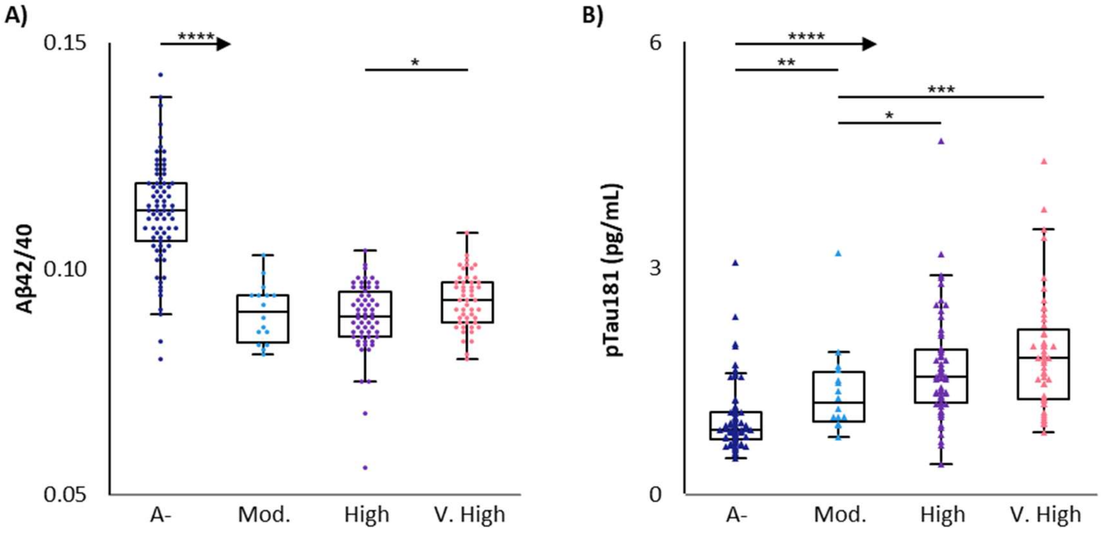

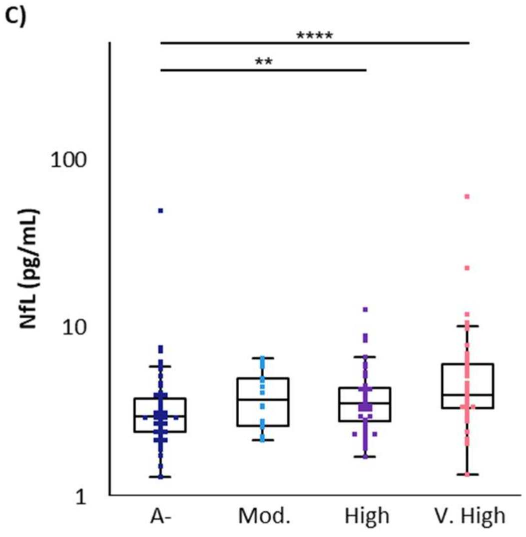

D)

| | Centiloid Values | | | |
|---|---|---|---|---|
| | A- (<15) | Mod. (26-50) | High (51-100) | V. High (>100) |
| Age (mean ± SD) | 73.8 ± 6.1 | 77.0 ± 7.6 | 73.9 ± 8.4 | 76.6 ± 6.8 |
| Sex (female/male) | 38 / 37 | 9 / 7 | 28 / 32 | 28 / 21 |
| APOE4 Status (- / +) | 65 / 10 | 8 / 8 | 27 / 33 | 16 / 33 |
| MMSE (mean ± SD) | 29.0 ± 1.3 | 28.4 ± 2.2 | 25.1 ± 3.7 | 22.7 ± 6.7 |
| n | 75 | 16 | 60 | 49 |

**Figure 4: NfL ROC Analysis with Respect to pTau181 Results**

NfL results obtained for AIBL samples were analyzed with respect to pTau181 status. This was defined using measurements and the cutoff (0.977 pg/mL) determined by ROC analysis of pTau181 results with respect to amyloid PET.

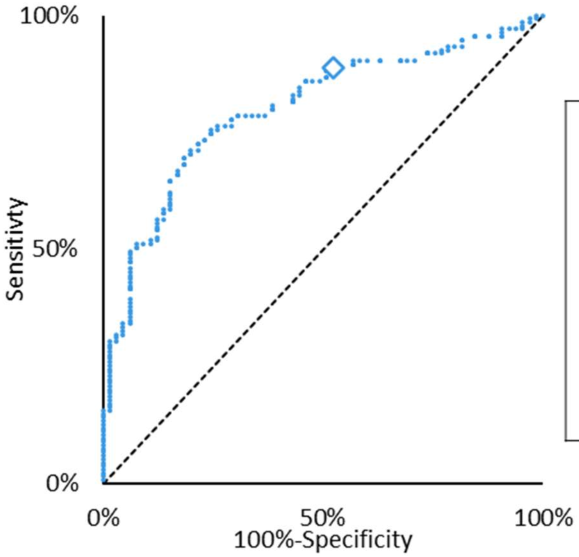

| | |
|---|---|
| AUC (95% CI) | 0.799 (0.74-0.86) |
| Cutoff (◇) | 2.62 pg/mL |
| Sensitivity | 88.9%% |
| Specificity | 47.7% |
| Efficiency | 75.5% |
| p-value | < 0.001 |
